# Exploration of wastewater surveillance for Monkeypox virus

**DOI:** 10.1101/2022.11.10.22282091

**Authors:** Edgard M Mejia, Nikho A Hizon, Codey E Dueck, Ravinder Lidder, Jade Daigle, Quinn Wonitowy, Nestor G Medina, Umar P Mohammed, Graham W Cox, David Safronetz, Mable Hagan, Jim Strong, Anil Nichani, Michael R Mulvey, Chand S Mangat

## Abstract

The sudden emergence and spread of Monkeypox in non-endemic parts of the world is currently not well understood. Infections are often mis-diagnosed and surveillance strategies are scarce. Wastewater-based surveillance (WBS) of human Monkeypox virus (MPXV) can help supplement our current clinical surveillance and mitigation efforts. WBS has shown to be an effective tool in monitoring the spread of other infectious pathogens, such as SARS-CoV-2 and its variants, and has helped guide public health actions. In this study, we describe how WBS can be used to detect MPXV in wastewater. We conducted WBS for MPXV in 22 wastewater treatment plants (WWTPs) over a period of 14 weeks. Nucleic acids were extracted using the MagAttract PowerMicrobiome DNA/RNA extraction kit. Three real-time qPCR assays were assessed for the detection of MPXV in wastewater. These included the G2R assays (G2R_WA and G2R_G) developed by the Centers for Disease Control and Prevention (CDC) in 2010, as well as an in-house-developed assay (G2R_NML). The G2R_G (generic) assay was designed to detect both the Congo and West African clades (re-named to Clades one and two, respectively) of viruses while the G2R_WA assay was designed to detect the West African clade (Clade one). The G2R_NML assay was designed using reference genomes of the 2022 MPXV outbreak. Our results show that all three assays have similar limits of detection and are all able to detect the presence of MPXV in wastewater. Following detection through real time qPCR, Sanger sequencing was performed on the resulting amplicon products, with the assembled contigs then undergoing analysis using nucleotide Basic Local Alignment Search Tool (BLAST). Due in part to the longer amplicon size of the G2R_NML assay, a significantly greater number of positive detections were identified as originating from MPXV compared to the CDC G2R assays. The ability to detect trace amounts of MPXV in wastewater as well as obtain Sanger sequence confirmation, has allowed for the successful surveillance of this virus in wastewater.

## Introduction

The WHO has declared the current Monkeypox (MPXV) outbreak as a Public Health Emergency of International Concern^1^. As of the writing of this report, 61,827 cases have been reported in 97 countries that are largely non-endemic for this disease^2^. MPXV was first identified in nonhuman primates in 1958 in a research setting ^3^, and the first human clinical case was identified in 1970 in the former province of Équateur, which is now known as the Democratic Republic of the Congo^4^. The first clinical case was later confirmed by serological investigation as MPXV^5^. Cases have since been generally well controlled and limited to the central west African region. Additional cases have been reported outside of this region in the UK, Israel and Singapore, with a notable 2003 outbreak linked to pet prairie dogs in the US^6^.

MPXV is a member of the Poxviridae family of DNA viruses, the Orthopoxviridae genus to which it belongs includes smallpox and whose waning global acquired immunity has been implicated in its spread^7,8^. Small mammals and primates are implicated as the hosts for MPXV and the natural reservoir is undefined^9^. MPXV has been detected in pox lesions, the upper respiratory tract, blood, urine, feces, and semen in infected individuals linked to the 2022 outbreak^10,11^.

The virus contains a 197 kb double stranded DNA genome packaged into a 200 × 250 nm virion that is described as “brick-like”^7,12^. MPXV is currently divided into three clades^12,13^; clade 1 (formerly Congo Basin clade) is typified by increased transmissibility and a higher case fatality ratio^14^. Clade 2 (formerly West African clade) tends to cause a milder disease and exhibits reduced transmissibility^15^. Isolates from the recent 2017 to 2019 outbreaks form the foundation for clade 3 and include cases from the on-going 2022 multi-country outbreak.

Molecular detection of MPXV by PCR has been elaborated in several studies describing assays that are genus, species, or clade specific^16–20^. The National Microbiology Laboratory, Canada’s national public health high-containment facility, supported clinical diagnosis of MPXV via four diagnostic assays; the B6R^16^ assay and G2R_G^18^ assays are species-specific assays directed at the envelope protein and TNF receptor, respectively. The G2R_WA^18^ assay is also directed at the TNF receptor gene and is specific to the clade 2 and 3, while the C3L^18^ assay is directed at the complement binding protein and is specific to the clade 1.

Limited testing, a wide range of disease presentation and stigmatized transmission amongst men who have sex with men (MSM)^10,11,21,22^ has likely increased the global spread of MPXV and alternate strategies are required to monitor the global spread of MPXV to prioritize infection control practices. Since the first detection of SARS-CoV-2 in sewage^23^, wastewater-based epidemiology has been an effective means to provide early-warning and trending of COVID-19 in congregant living settings, small and large communities that have been communicated in hundreds of articles too numerous to provide here. The multiple shedding routes of MPXV, including stool shedding, present an opportunity to apply WBS to current MPXV outbreak. In this work, we compare the performance of the previously described G2R_WA and G2R_G assay described by Li and co-workers^18^ with a novel assay named G2R_NML. We then apply this assay to demonstrate through limited surveillance across various Canadian cities that signal derived from both these assays provide public health value by providing forewarning on the appearance of clinical cases.

## Materials and Methods

### Sewage sample collection

Influent 24 hr-composite wastewater samples were collected from 10 anonymized Canadian cities with collection dates ranging from June 28 to September 30th, 2022. Some 22 wastewater treatment plants were included in the study, where a major metropolitan center in 9 of 10 Canada’s provinces, one province was represented by two cities. Population coverages for each province ranged between 21% to 57% (median coverage of 22 %), some 23% of all Canadians were represented in this study. Typically for most sites, two sewage samples were processed weekly from all locations. Flow data was also collected from these six communities. Samples were collected in sterile 500 mL polyethylene terephthalate (PET) bottles and were shipped to the National Microbiology Laboratory at 4°C. Samples were stored at 4°C in the dark until and processed within two days of receipt (Figure 1).

**Figure 1.**
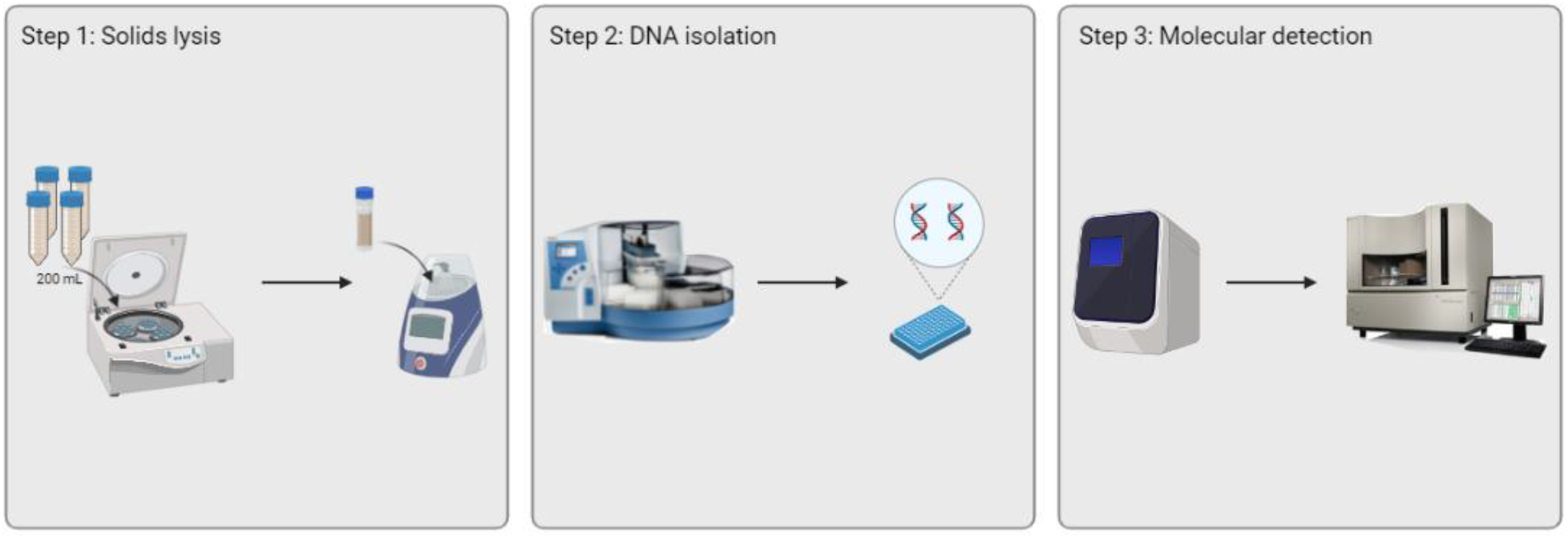
Workflow to detect MPXV in wastewater. Step 1 includes centrifuging 200 mL of sewage to obtain a solids pellet that is bead-beaten in a tube containing lysis buffer and silica-zirconium beads. Step 2 depicts DNA isolation on the KingFisher Flex using the Qiagen MagAttract PowerMicrobiome kit. Step 3 utilizes qPCR to detect and quantify MPXV targets. Following qPCR, the amplicons are Sanger sequence confirmed.

### Sample processing and nucleic acid isolation

We had initially attempted to survey for the MPXV signatures in wastewater in two epicenters of the infections over the period of May through June 2022 and were unable to garner any signal (Data not shown). Briefly, the assay conditions were to extract either the insoluble material evolved from centrifugation of 30 mL of raw wastewater, or, through concentration of 15 mL of wastewater using a 10 kDa centrifugal filter device. In both cases nucleic acid was extracted using the Roche MP96 extraction platform using the DNA and Viral NA Large Volume extraction kit and detection using the G2R_WA assay described by Li and co-workers. Only when we increased the processed volume to 200 mL and used the Qiagen PowerMicrobiome extraction kit were we able to detect MPXV in Canadian wastewater. This change was inspired by a social media post from Dr. Alexandria Boehm’s group reporting on positive detections of MPXV in California and most recently reported as a pre-print using a method targeting the settled solids of wastewater. The group has since published a pre-print article^24^ that states that MPXV is present 10^3^ times more in the insoluble vs soluble fraction of wastewater, which is supported by the work presented here.

A 200 mL sample of well-mixed primary influent was centrifuged for 30 min at 12,000 x g at 4°C to yield a pellet mass that was used for subsequent processing. Nucleic acids were isolated from the wastewater solids using the MagAttract PowerMicrobiome DNA/RNA extraction kit (Qiagen, Hilden, Germany) on the Kingfisher Flex (ThermoFisher Scientific, Waltham, MA). The pellets were re-suspended in 650 μL of heated MBL solution containing 2.5% 2-mercaptoethanol and transferred to a 2 mL PowerBead Pro tube (Qiagen, Hilden, Germany). Individual bead beating tubes were chosen over the plate format to improve lysis and recovery^25^. The pellet-bead mixture was processed using a Bead Mill 24 Homogenizer (Fisher Scientific, Ottawa, ON) at 6 m/s for three 60 s cycles with a 10 s rest. Manufacturers’ recommendations were followed after sample lysis with the addition of 2 uL of ≥ 9 mg/mL carrier RNA to each well (Sigma-Aldrich, Oakville, ON). The median effective sample volume was 128.6 mL due to lysate produced in excess of the maximum loading volume (450 uL).

### Real-time qPCR assay development and validation

Primers and probes (Table 1) for the G2R_NML real-time qPCR assay were developed *in silico* using PrimerQuest™ Tool from Integrated DNA Technologies (IDT). The target gene for this assay is the MPXV G2R gene (1050 bp in length) on the MPXVgp190 locus of the Clade 2 viral genome. Using GenBank, we obtained the first complete MPXV genome sequence (MPXV_USA_2022_MA001; accession ON563414) of the current 2022 MPXV outbreak and used it as a reference genome to develop the G2R_NML assay. To determine assay specificity, the primers and probes were run through NCBI BLAST. Additionally, utilizing the GISAID genome database, we obtained supplementary MPXV genomes from the current outbreak and used them as reference genomes to determine the specificity (*in silico*) of the G2R_NML assay. Additionally, we tested the MPXV qPCR assays developed by Li et al^18^ and compared their performance to the G2R_NML assay.

**Table 1:**
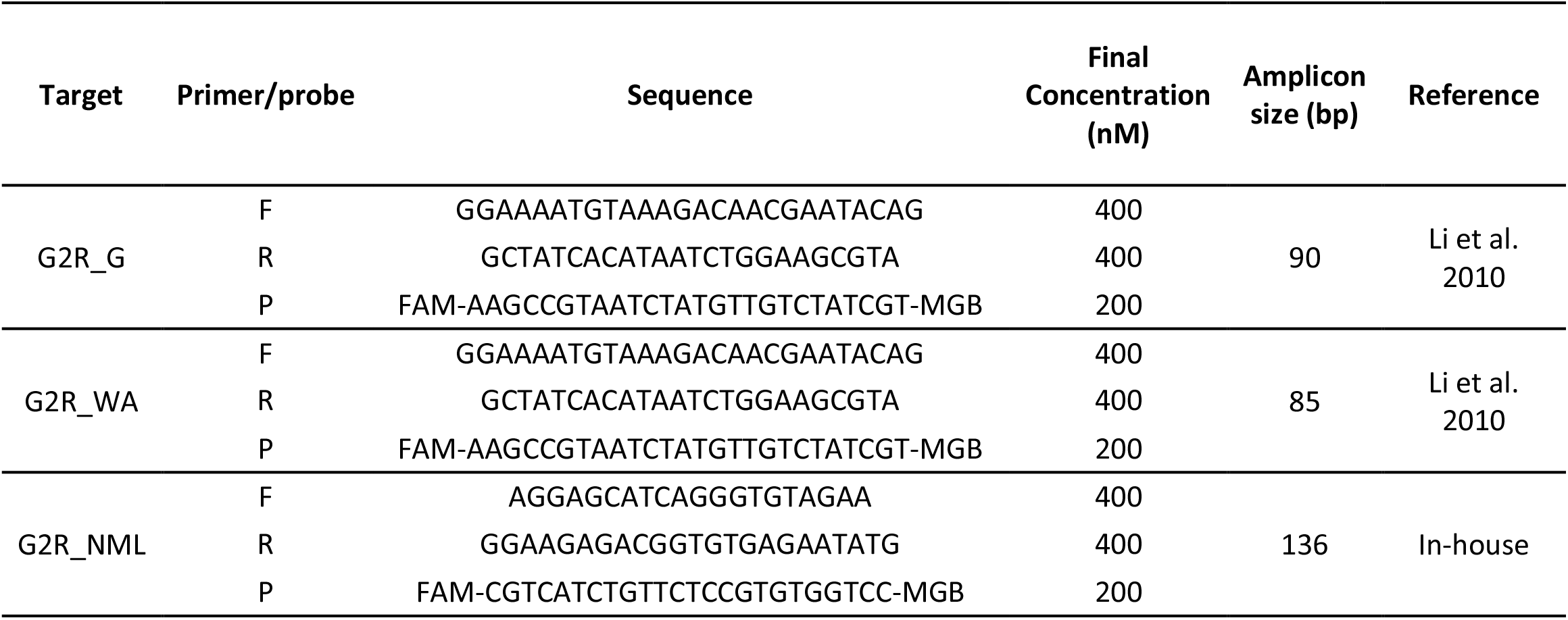
G2R_G, G2R_WA, G2R_NML primers and probes sequences, final concentrations and amplicon lengths.

### Real-time qPCR

Primer and probe sequences are shown in Table 1. Assay conditions for the three MPXV qPCR assays were as follows: qPCR was performed in a reaction volume of 20 μL, consisting of 5 μL QuantiNova Multiplex PCR Kit (Qiagen, Hilden, Germany), 400 nM (G2R_G, G2R_WA) and 300 nM (G2R_NML) final concentration of each respective primer set (ThermoFisher Scientific); 200 nM (G2R_G, G2R_WA) and 250 nM (G2R_NML)) of each probe (ThermoFisher Scientific); and 5 μL of template DNA and Invitrogen nuclease-free H_2_O (ThermoFisher Scientific). PCR amplification and detection of amplification products was performed on a QuantStudio 5 Real-time PCR instrument (ThermoFisher Scientific). Thermal cycling conditions were as follows: polymerase activation at 95°C for 2 min, followed by 40 cycles of 95°C for 5 s and 60°C for 30 s. Each real-time PCR was performed in triplicate with the appropriate non-template controls and positive controls. Results were considered positive if they had a Ct value <40 (25). A standard curve was prepared for each target by preparing a five-point serial dilution (50,000 – 5 cp/rxn) of linearized plasmid containing the plasmid sequence (Thermofisher) (Table 2) with nuclease-free water and qPCR was run in triplicate for each point.

**Table 2:**
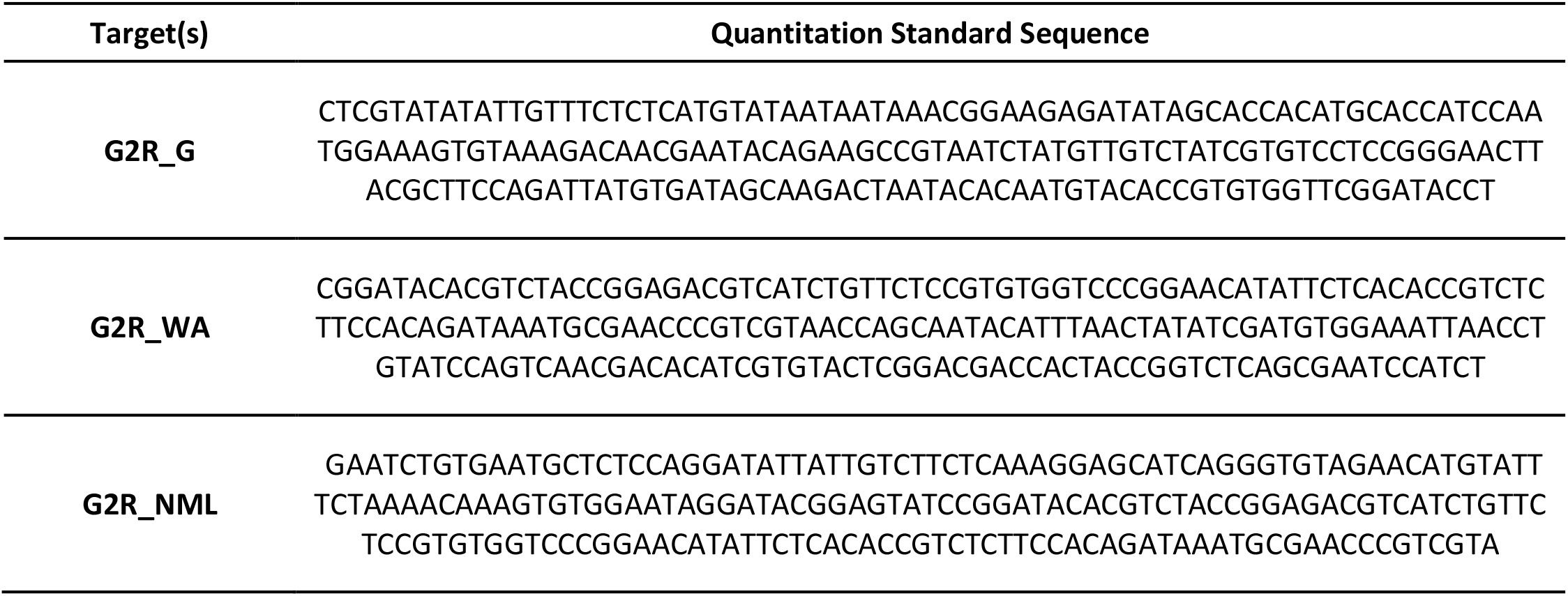
MPXV 200bp DNA fragments for G2R_G, G2R_WA, and G2R_NML targets.

### Assay Sensitivity

Standard reference material in the form of synthetic DNA fragments were created (Thermofisher Scientific) to support assay development and evaluation and were designed against G2R_G, G2R_WA, and G2R_NML target regions (Table 2). The three MPXV standards consisted of 200 bp regions unique to the primer probe sets, which were inserted into individual pMA-RQ (AmpR) GeneArt cloning vectors (Thermofisher Scientific). These plasmids were linearized using a high-fidelity ScaI restriction enzyme (New England BioLabs) and were purified using the QIAquick PCR Purification Kit (Qiagen). TapeStation DNA ScreenTape (Agilent) was run on the plasmid constructs to confirm linearity. Standards were quantified using the Absolute Q™ Digital PCR System (ThermoFisher) following manufacturers’ recommendations.

Assay limits of detection (ALOD)^26^ were determined by measuring the relationship between positivity rates of standardized qPCR reactions across a 13 point serial dilution of standard reference material (as described above, see Table 2) from 45 copies/reaction (cp/rxn) to 0.1 cp/rxn. For each reference material concentration, 20 replicates were performed and a Ct value of < 40 was deemed to be a positive detection. A Probit model was fit to positivity rates for each assay and the ALOD was determined from where the regression model intersected with a 95% positivity rate. The ALOD confidence interval was estimated from where the regression model’s confidence interval intersected at the 95% assay positivity. The Assay Limit of Quantification (ALOQ)^26^ was determined by measuring the relationship between the coefficient of variance of standardized qPCR reactions across an 11 point serial dilution of standard reference material from 45 to 3.95 cp/rxn. Reaction positivity and standard reference material are as described above. Coefficient of variation (CV) was calculated according to equation below.

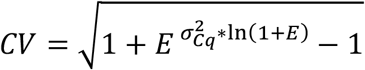

Where E is the PCR efficiency as described above, 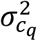 is the Cq standard deviation of the cycle thresholds corresponding to a specific concentration. The CV for each assay was fit to a linear regression model against the log_10_ cp/rxn and the determined CV for each reaction. The ALOQ was where the line of best fit intersected with a CV of 35%.

The sample limit of detection^26^ (SLOD) and sample limit of quantification^26^ (SLOQ) was calculated using the median effective sample volume, template volume, and extraction volume (100 uL) – see also^27^.

### Sanger sequencing of real-time qPCR products

Triplicate qPCR amplification products were pooled into a total volume of 60 μL. Pooled amplicons were purified using Ampure XP Reagent (Beckman Coulter catalog number: A63881) following the manufacturers’ instructions, with a modified elution volume of 20 μL. A volume of 1 μL of purified amplicon was Sanger-sequenced using BigDye-TerminatorTM v3.1 Cycle Sequencing Reaction Kit on a 3730 xl Genetic Analyzer (Applied Biosystems, Foster City, CA, USA) following the manufacturer’s instructions. PCR sequences were aligned using the Seqman Pro15 assembly program as part of the DNASTAR Lasergene 15 software suite. Assembled contigs were analyzed with nucleotide Basic Alignment Search Tool (BLAST) assigned to megablast optimization for highly similar sequences. Query results were analyzed for top matches related to Monkeypox specific query results with equal to or greater than 95% bp query coverage and equal to or greater than 95% identification.

### Clinical case data

First case reported dates were gathered from an online web-portal https://www.canada.ca/en/public-health/services/diseases/monkeypox.html.

## Results

To augment the complement of qPCR assays to address the emerging MPXV outbreak, a qPCR assay was designed against the TNF receptor gene located within the 5’ and 3’ terminal repeats of the virus, named G2R_NML. The previously published G2R_WA assay targets this same loci^18^ and indeed the reverse primer of the G2R_NML and the forward primer of the G2R_WA overlap. Figure 2a and 2b depict alignments of all of these assays’ primer and probe sequences against a panel of Clade 2 genomes drawn from recently sequenced genomes of the 2022 outbreak. Seven nucleotides of the 5’ probe of the previously published G2R_WA assay are non-complementary when aligned to a panel of Clade 1 genomes, as this design feature affords the discriminatory ability of this assay against Clade 1 and Clade 2 variants (Figure 2C). Figure 2d shows the alignment of the G2R_G assay against Clade 1 genomes, which shows its ability to also detect this MPXV clade. The amplicon sizes of the G2R_NML and G2R_WA assays are 136bp and 85bp, respectively (Table 1). The G2R_G assay from Li *et al* (18), which can detect both clade 1 and 2 variants, was also employed in this work.

**Figure 2.**
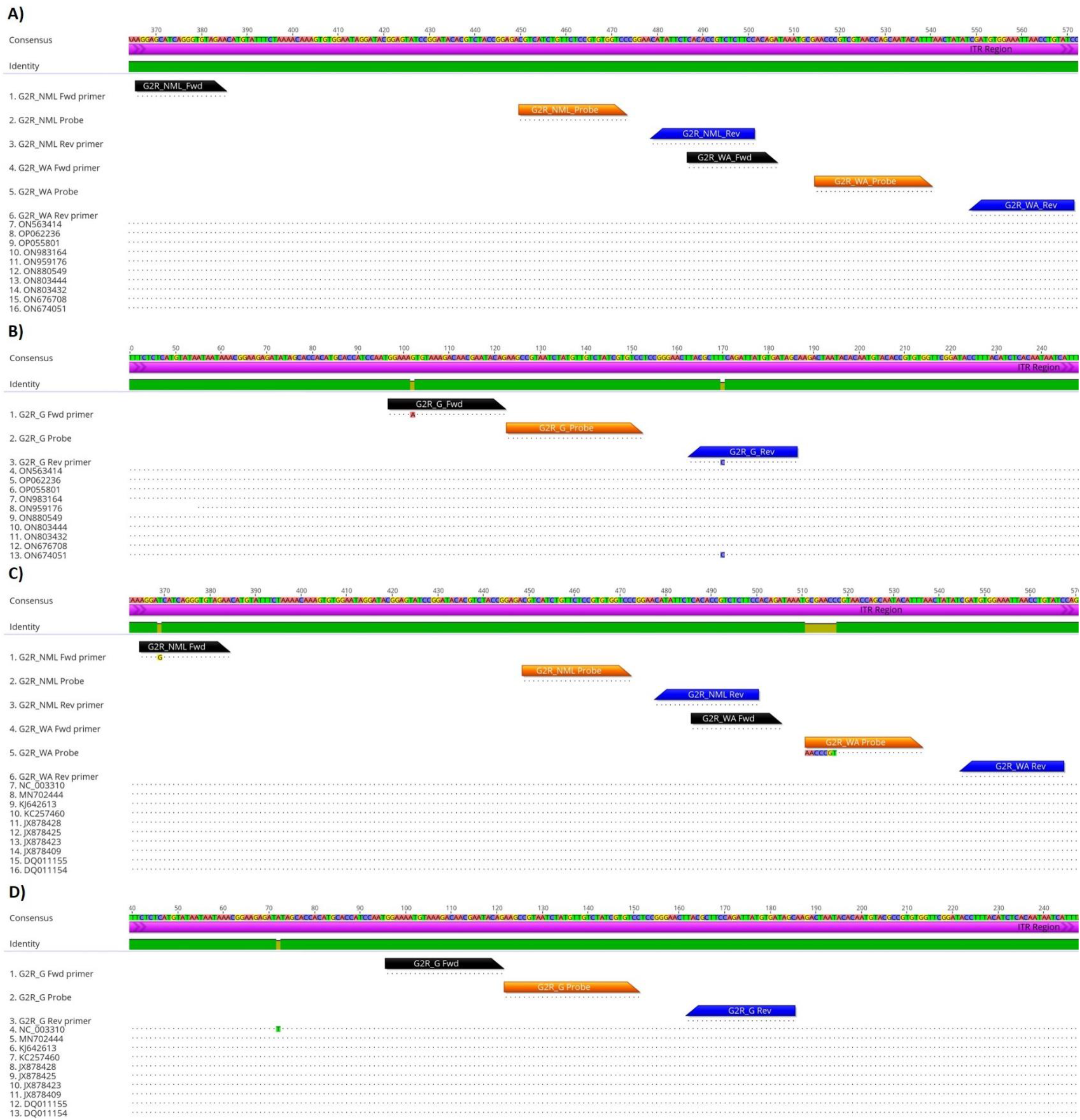
Alignment of MPXV assay primers and probes to reference Clade two and Clade one strains. **A)** Alignment of the G2R_NML, G2R_WA and **B)** G2R_G primers and probes to the TNF receptor gene from reference Clade two MPXV genomes. **C)** Alignment of the G2R_NML, G2R_WA and **D)** G2R_G primers and probes to the TNF receptor genes from reference Clade one MPXV genomes. Identity graph illustrates pairwise-identity homology (green = match, light brown = mis-match). Purple graph illustrates the inverted terminal repeat (ITR) region. Image created using Geneious Prime^®^ version 2022.0.2.

The log-linear slope, intercept, linear correlation, and PCR efficiencies of all of the above assays is described in Table 3. The slopes for each assay ranged from −3.43 to −3.55, and delivered PCR efficiencies of 91% to 96%. The correlation coefficient across all assays ranged from 0.9956 to 0.9976. Intercepts across all assays ranged between 37.30 and 38.13, and none of the no-template control runs for each assay registered as positive.

**Table 3:** Real time qPCR assay parameters for G2R_G, G2R_WA, and G2R_NML

| Target | Slope | Intercept | RSQ | Efficiency |
| --- | --- | --- | --- | --- |
| G2R_G | -3.55 | 38.13 | 0.9956 | 91% |
| G2R_WA | -3.49 | 37.62 | 0.9959 | 93% |
| G2R_NML | -3.43 | 37.30 | 0.9976 | 96% |

The ALOD (95% CI) for the G2R_G, G2R_WA and G2R_NML assays were 3.52 (2.47, 6.03), 3.47 (2.37, 6.28) and 3.48 (2.42, 6.03) cp/rxn, respectively (see Figure 3A-C, vertical black line). This very closely matches what Li et *al*^18^ found in their study where they report LOD values of ~3.5 and ~8.2 genomes from the G2R_G and G2R_WA assays, respectively. This implies a SLOD (95% CI) for the G2R_G, G2R_WA and G2R_NML assays were 0.55 (0.39, 0.94), 0.54 (0.37, 0.98) and 0.54 (0.38, 0.94) cp/mL. The ALOQ for the G2R_G, G2R_WA and G2R_NML assays were 17.2, 19.9 and 20.1 cp/rxn, respectively (see Figure 3D-F, vertical black line). The SLOQ for the G2R_G, G2R_WA and G2R_NML assays were 2.68, 3.11 and 3.14 cp/mL.

**Figure 3.**
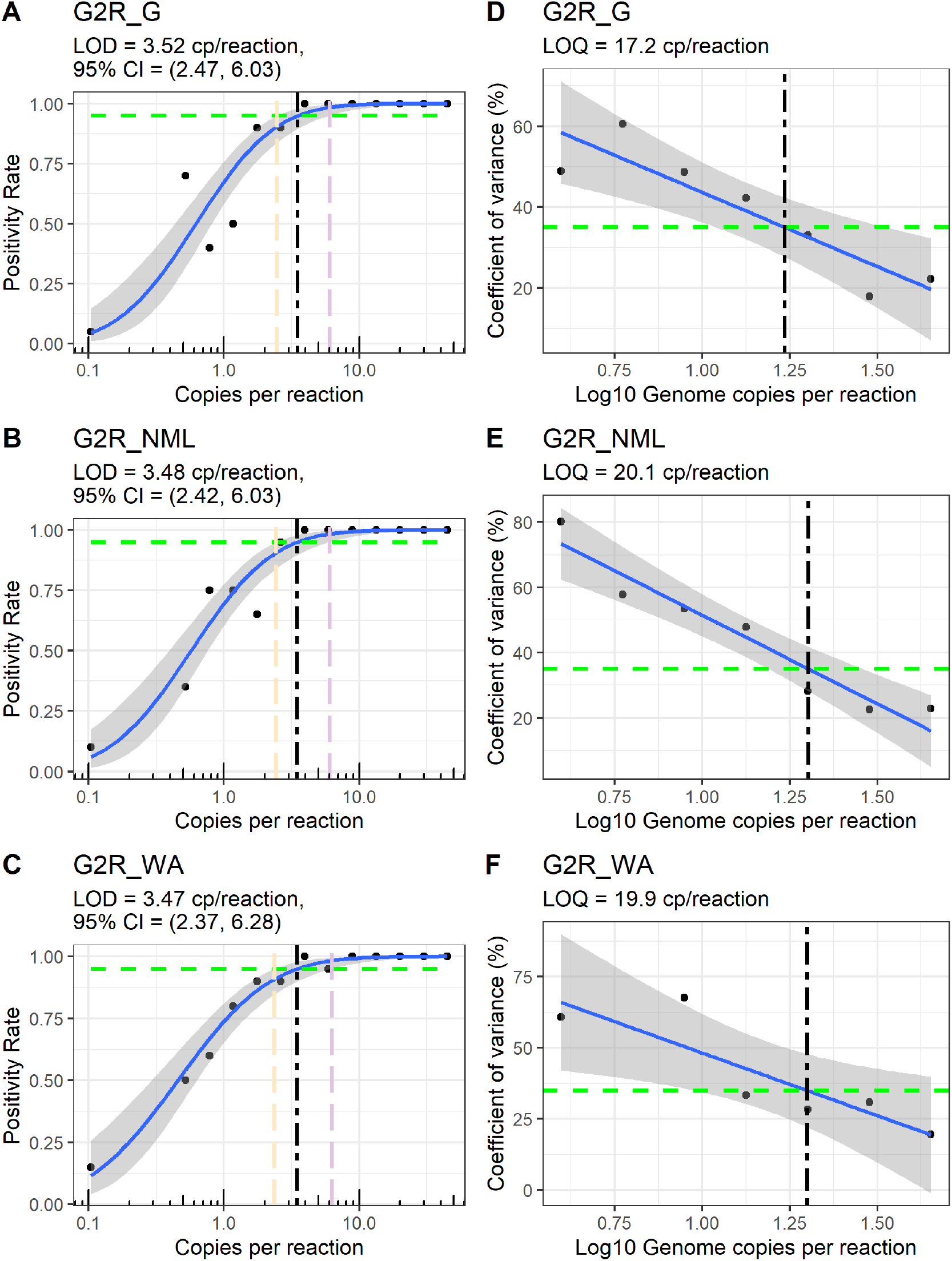
Limits of detection and quantification of G2R_G, G2R_NML, and G2R_WA assays. **(A-C)** Positivity rates of the 3 assays obtained from a 15 serial dilution were fitted to a Probit regression for the estimation of the LoD (solid blue line). The 95% confidence interval from the regression fit (grey band) allows estimating the 95% confidence interval for the LoD (orange and purple dashed line). **(D-F)** Coefficient of variance of the 3 assays obtained from a 15 serial dilution were fitted to a linear regression for the estimation of the LoQ (solid blue line).

For each assay, triplicates from qPCR positive wastewater samples were pooled and Sanger sequenced, then aligned to Monkeypox via BLAST analysis. The sequence confirmation rates for the G2R_G, G2R_WA, and G2R_NML assays were 16%, 22%, and 76%, respectively (χ^2^ p-value = 89.9 (3.07×10^−20^), Table 4). Negative result denotes a false positive or a sequence fail.

**Table 4:** Proportion of positive real time qPCR signals that were confirmatory for MPXV via Sanger sequence analysis.

| Monkeypox<br>Sequence<br>Match | G2R_G (%) | G2R_WA (%) | G2R_NML (%) | X<sup>2</sup> (p-value) |
| --- | --- | --- | --- | --- |
| Negative | 82 (84) | 98 (78) | 27 (26) | 89.9 (3.07×10<sup>-20</sup>) |
| Positive | 16 (16) | 28 (22) | 76 (74) |  |

The viral load from wastewater samples drawn from all communities tested in this work were determined using the G2R_G, G2R_WA and G2R_NML assays in triplicate. The distribution of the censored geometric mean of replicate viral loads for all samples collected from July 7^th^ to August 25^th^ is presented in Figure 4A-left. The sample mean for the G2R_G, G2R_WA and G2R_NML assays were 0.52, 0.45, and 0.51 cp/mL, respectively. These values were supported by Ct values of 36.3, 36.1 and 35.6 for the G2R_G, G2R_WA and G2R_NML assays, respectively (Figure 4A-right). Spearman’s correlations and a pair-wise comparison for all assays is presented in Figure 4 B-D. The correlations were as follows G2R_WA:G2R_NML was 0.717, G2R_G:G2R_WA was 0.754, and G2R_G:G2R_NML was 0.755.

**Figure 4.**
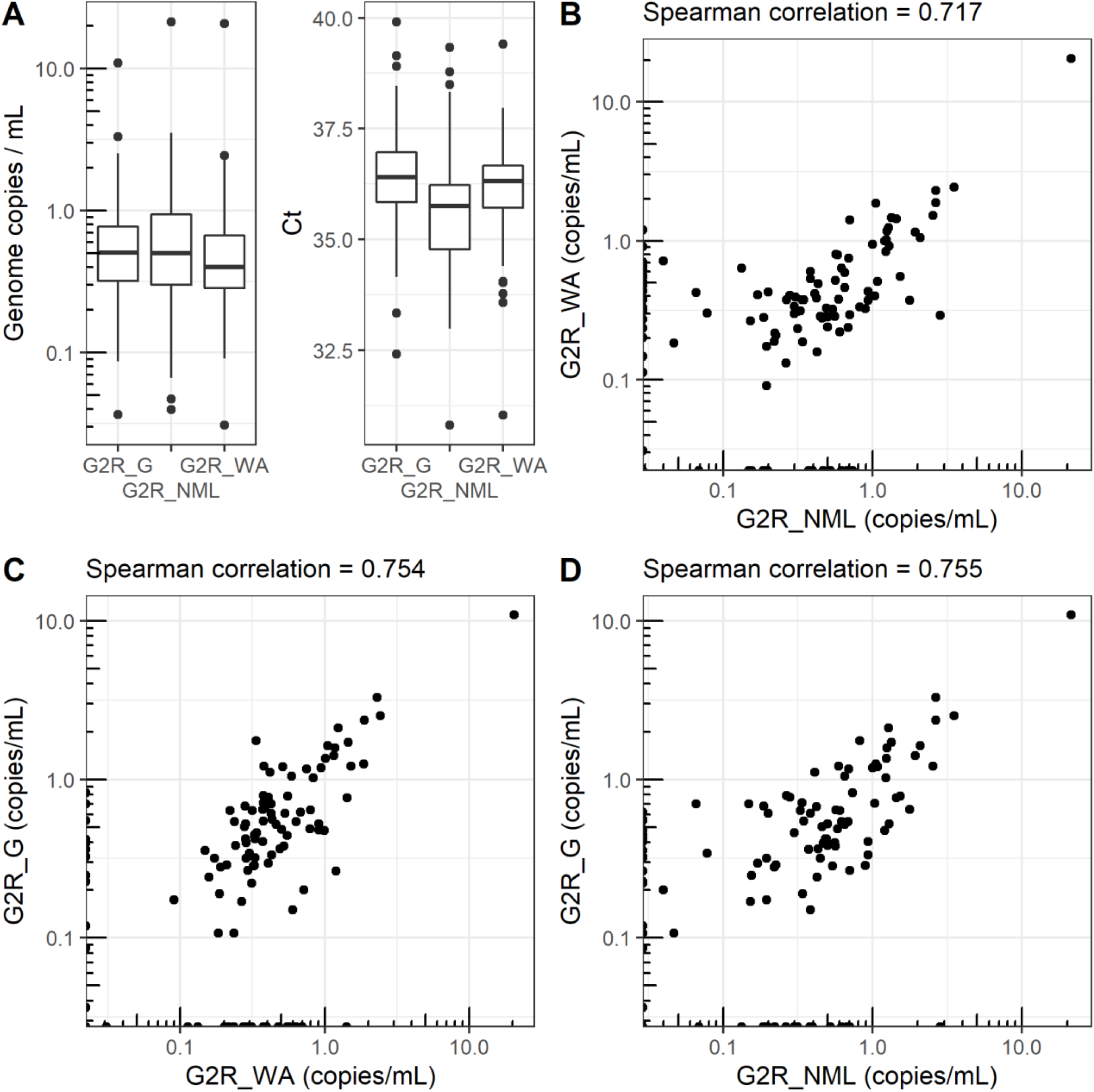
Performance of MPXV qPCR assays. **(A-left)** Viral load distributions and supporting **(A-right)** Cts across the indicated assays for all communities tested is presented as a box and whisker box plot; the edges of the box of indicate the upper and lower quartiles, the whiskers extend to the values that are no further than 1.5 times the interquartile range, observations outside the whisker extent are represented by a dot. **(B-D)** Pair-wise comparison of log-transformed unadjusted viral loads for assays as indicated on the axes, non-detects are presented on their respective axis line. Spearman correlation for each pair-wise comparison are indicated above each plot.

All positive samples were submitted for Sanger sequencing. As described in Table 4, the % positivity rate for G2R_G, G2R_WA and G2R_NML, was 16%, 28% and 76%.

Comparison of assay positivity across all three assays across all sites tested during the study period are represented in Figure 5. A major metropolitan center in nearly each Canadian province was tested over the 14 week study period. For one province, two city sites were tested (Figure 5 – “Province D”). If any wastewater treatment plant registered as positive for the week in a province, the entire week was marked as positive (Figure 5 – red boxes). For 5 provinces the first clinical detection was outside the study period for this work. For provinces D and H wastewater detection of MPXV was 7 and 3 weeks prior to clinical detection, respectively. For province G and I wastewater detection of MPXV was 5 and 1 weeks following the first clinical detection, respectively.

**Figure 5.**
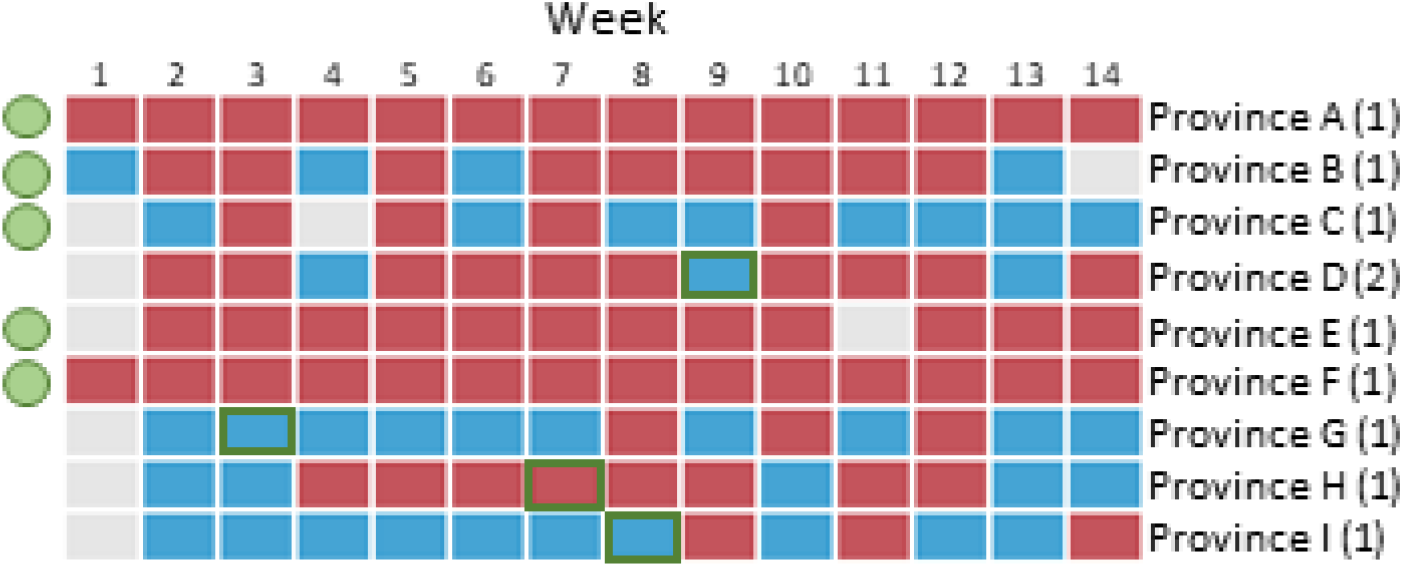
Comparison of MPXV detections in wastewater across Canada and first clinical detections. Each row of the diagram represents surveillance in a major metropolitan centre for 9 Canadian provinces, except for province D where two cities represent the province. Each box in the matrix diagram represents a week of surveillance; for each week wastewater samples were tested across the three PCR assays described in this work, a positive detection by any assay is represented in red, a negative detections across all assays are represented in blue, and grey boxes indicate no sample was received from the site. The first clinical detection reported at the provincial for each site during the study period are indicated by a green box, where first clinical detection was outside the bounds of this work are indicated by a green circle to the left of the diagram.

## Discussion

Currently, information on the fecal shedding of MPXV is limited. Adler *et al* have shown that MPXV DNA can be found in the upper respiratory tract, blood and urine of infected patients^10^, while Antinori *et al* have shown that MPXV DNA can be found in seminal fluid and feces as well^11^. Its widespread bodily secretion and excretion suggest the possibility that this virus can be tracked using WBS. With limited capacity for clinical testing and the stigma often associated with this illness, WBS can act as a complimentary surveillance tool not only to enhance surveillance efforts but also to provide a fulsome picture of the extent of MPX activity.

In this work, we demonstrate that the G2R_NML assay is complementary to both G2R_G and G2R_WA from Li and co-workers^18^ for detection of MPXV in wastewater. All assays have similar LODs and LOQs, and correlate well with each other as demonstrated by the high Spearman correlation. Most of the detections in this work were below the SLOD and SLOQ, despite a clear correlation across all assays. For this reason we favored interpretation of the data as a binary signal driven by assay positivity. MPXV wastewater signals were a leading indicator of reported clinical cases at the provincial level at provincial sites D and H and closely correlated with clinical detection at provincial site I. At provincial site G wastewater lagged considerably from clinical detection, although the lack of geo-location of clinical signal hampers precise interpretation. These data could be supportive of informing the allocation of resources such as vaccines to vulnerable populations with the areas of detection.

The specific advantage of NML-developed assay is that amplicons are more easily confirmed by Sanger sequencing. Likely, because the design of this assay has a slightly larger amplicon (136 vs 85/90 bp). This increase does not impact the qPCR efficiency, and the ALOD and ALOQ are comparable to the CDC assays. Amplicons derived from environmental surveillance are a challenge to sequence due to the high-diversity of DNA in samples, which can lead to off-target amplification that inhibit sequencing efforts. This is made doubly difficult as signals tend to be at the technical limitations of qPCR. Sub-cloning amplicons is a viable but cumbersome route to garnering sequence confirmation. Since the current Monkeypox outbreak is an emerging and high-consequence issue for public health decision makers, timely access to sequence confirmation of emerging signals promotes uptake of this novel data stream. A recent study conducted by de Jonge *et al*^28^ describes an elegant approach to confirm the qPCR signals obtained from MPXV detection in wastewater. In brief, a semi-nested qPCR was used to generate amplicons using the forward primer of the G2R_G and the reverse primer of the G2R_WA assays. The product was then put through another round of amplification using the G2R_G probe and G2R_WA reverse primer to generate an amplicon that could be Sanger sequenced^28^. Additionally, the distribution of Cts for the G2R_NML assay is lower than the G2R_G and G2R_WA assays, suggesting that the performance of this assay is at least sensitive and perhaps more sensitive than the above mentioned assays. In sum, we feel that the G2R_NML assay can act as a complement to developing MPXV wastewater surveillance systems.

Finally, for COVID-19 WBS, many groups include two targets as part of their assay regiment to mitigate the inherent variable nature of WBS and the risk of mutation. Given that there are indications that the recent Monkeypox outbreak could be driven in-part by a higher mutational frequency^12^, a second assay as part of a WBS surveillance program is a means to mitigating loss of sensitivity due to mutation. The CDC assays afford high sensitivity, yet the route to confirmation has proven challenging. The G2R_NML assay could be a useful tool when this confirmation is required.

The Boehm group and others^29,30^ have been advocates for WBS assay designs that extract genetic material from either the settled solids from primary influent/raw sewage or through high-solids samples sampled from the primary clarifier. The high prevalence and attack rate of SARS-CoV-2 have likely allowed for smaller process volumes to support COVID-19 surveillance that is minimally tracking clinical cases and hospitalizations. However, the current MPXV outbreak shows that higher process volumes that are directed to the insoluble fractions are an operational necessity. To re-orient WBS systems to the process of high-solids contents is difficult as larger sample volumes are required to garner adequate material for processing, which can complicate sample collection, increase overhead of laboratory management and reduce throughput over smaller sample volumes. Standard methods to concentrate solid material at the point of sampling are a potential solution to alleviate the logistical burden of shipping large volumes of wastewater to support a central-laboratory model of WS.

To further develop MPXV WBS, several open questions should be answered; the stability of the virus across temperature and time should be performed, as it was for SARS-CoV-2. This will help define hold-times that ultimately limit/define the surveillance structure. Furthermore, an epidemiological useful scheme to track the mutations profile through metagenomics-like methods is required. As MPXV is a large virus this will likely be an onerous task. However, focusing on the immunologically relevant regions may be a route to an efficient and informative design, this is supported by observations that a signature change in MPXV genome over this recent outbreak have been changes to surface antigens present in the virus.

Future work to test the stability of the virus in wastewater is still required. Additionally, while we made every attempt to make the G2R_NML assay specific to MPXV by in silico analysis, cross-reactivity studies involving other orthopoxvirus targets are still required. Also, further comparisons between clinical and wastewater signal trends are needed. Correspondence to clinical surveillance would be useful in further establishing the confidence of this assay, however, stigmatization could erode this correspondence. Case reported dates may be temporally separated from peak fecal shedding which is likely at the onset of symptomology. This necessitates careful follow-up studies as correspondence between clinical and wastewater signals requires both a temporal and spatial element.

The WBS for MPXV will enable jurisdictions to monitor the virus in their sewershed even when no clinical cases are known to be present. This will help make important resource allocation decisions in the current context when the system is overwhelmed with the COVID-19 response. With no MPXV signal in wastewater, other public health priorities can be supported with more confidence.

## Conclusions

The cause of the 2022 MPXV worldwide outbreak is not well understood. However, it highlights the fact that additional surveillance tools are necessary to better track and help contain its spread. This work suggests that WBS can be used as a viable surveillance strategy for the current MPXV outbreak.

## Data Availability

All data produced in the present study are available upon reasonable request to the authors

## Acknowledgements

We acknowledge the work of Jason Agasid for the quantification of the standard material. We acknowledge our collaboration with Statistics Canada for joint collection of wastewater through Canadian Wastewater Survey. We thank the municipalities for providing wastewater and participating Public Health authorities.

## Notes

### Competing Interest Statement

The authors have declared no competing interest.

### Funding Statement

This study was funded by internal funds (Government of Canada)

